# Effects of dyad motor practice on proprioceptive function

**DOI:** 10.1101/2025.04.25.25326388

**Authors:** Leoni Viola Winter, Jacquelyn VL Sertic, Jürgen Konczak

## Abstract

Dyad practice of complex motor skills, characterized by two learners alternating between physical and observational practice, can yield better motor outcomes and reduce practice time compared to physical practice alone. It is unknown if the superior effects of dyad practice on motor learning extend to proprioceptive learning.

Forty-two healthy participants (18–35 years) were randomized into three groups (n=14 each): Dyad practice, physical practice with rest (PP-rest), and physical practice without rest (PP-no rest). Participants practiced a 2 degree-of-freedom gamified wrist movement task for 20 minutes using a custom-made wrist robotic device. Wrist position sense acuity was assessed before (baseline) and 24 hours after the end of training (retention), using the Just-noticeable-difference (JND) threshold and Uncertainty.

Only the PP-no rest group exhibited significantly lower JND thresholds at retention compared to baseline (*t*(13)=2.44; *p*= 0.03, Hedge’s *g*=0.70). There were no differences in position sense Uncertainty within or between groups.

Dyad practice may yield superior gains in motor performance, but this did not translate into comparable gains in proprioceptive acuity. A possible explanation for these findings is that the recruitment of explicit motor learning mechanisms during dyad motor skill practice does not enhance the implicit learning mechanisms underlying proprioceptive learning.

**Highlights:**

- Dyad practice (DP) may yield superior motor gains compared to physical practice
- DP does not yield superior proprioceptive gains compared to physical practice
- Intensive physical practice yields the largest gains in position sense acuity

## Introduction

Motor and proprioceptive learning are closely coupled, such that motor learning enhances proprioceptive function and vice versa (Borich et al., 2015; Ostry et al., 2010). Numerous studies show that sensorimotor training induces both motor performance and proprioceptive gains (for reviews see Aman et al., 2015; Seo et al., 2023; Winter et al., 2022). For example, training of a visuomotor task in a wrist robotic environment can improve performance as well as proprioceptive acuity in neurologically healthy people and those presenting with chronic stroke or Parkinson’s disease (Elangovan et al., 2017, 2018, 2019).

The reciprocal neuroanatomical connections between motor and somatosensory cortical areas are well delineated (Ostry & Gribble, 2016). Substantial empirical evidence shows that motor learning leads to altered functional connectivity beyond motor cortical areas. For example, learning to adapt to unknown force fields while executing goal-directed arm movements leads to electrophysiological changes in networks involved in the processing of somatosensory afferents such as the somatosensory cortex (Vahdat et al., 2011). Conversely, applying continuous theta-burst magnetic stimulation before force-field training to disrupt activity in the primary somatosensory cortex impairs adaptive motor learning and retention (Darainy et al., 2023). In addition to changes in somatosensory networks following motor adaptation, research also shows that sensory learning induces changes in the motor areas of the brain, including increased motor evoked potentials and increased activity in the motor cortex (Ostry & Gribble, 2016).

Dyad practice is a motor learning paradigm in which individuals train in pairs (dyads) and alternate between physical training and observing the practice of the other member of the dyad. Empirical research shows that dyads perform at a similar or superior level compared to individuals who only practice physically in a variety of complex motor tasks, including visuomotor tasks such as flight simulation (Arthur et al., 1997; Shebilske et al., 1998) and a car racing game (Winter et al., 2024), microsurgery (Bjerrum et al., 2014), cup stacking (Granados & Wulf, 2007), balance training (Shea et al., 1999), trajectory learning (Panzer et al., 2019), and ultrasound simulation (Tolsgaard et al., 2015). Notably, dyad practice has received significant attention as a tool in motor task learning that requires costly equipment – such as microsurgery (Bjerrum et al., 2014), flight simulation (Shebilske et al., 1998), and simulation-based ultrasound training (Tolsgaard et al., 2015) – making dyad practice a valuable technique for complex motor skill learning in environments with scarce resources. The alternation between physical practice and observation may afford unique learning opportunities by accessing three distinct processes: 1) motor control processes related to movement execution (e.g. error correction, parameterization), 2) cognitive processes related to the movement task (e.g. error detection, information about correct movement pattern) (Shea et al., 2000), and 3) cognitive processes unique to action observation (e.g., detecting effective learning strategies and techniques (Panzer et al., 2019).

We recently assessed the effects of dyad practice on motor performance in a gamified exercise using a wrist robotic device (Winter et al., 2024). The results show that the additional observational practice in the dyad practice group led to superior motor performance immediately after training and after a 24-hour retention period compared to groups that practiced physically only. However, the effects of dyad practice on proprioceptive learning are unknown. Considering the motor skill learning benefits of dyad practice and given that proprioceptive and motor learning mechanisms are closely linked, the present study aimed to determine the effects of dyad practice on proprioceptive function. This report extends previous data on the effects of dyad practice on motor performance in a controlled, wrist-robotic environment (Winter et al., 2024) by providing data on the learning-related proprioceptive changes in wrist position sense.

## Methods

### Participants

Forty-two individuals (32 female, age 27.1 (SD 1.6) years) participated in the study. Inclusion criteria were: (1) age between 18-35 years, (2) right handedness, and (3) fluency in the English language. Individuals were excluded if they had any of the following: (1) uncorrected visual impairment, (2) neurological or physical impairment of the right upper extremity, or (3) speech pathology that impairs ability to communicate. All participants provided a verbal medical history to determine eligibility. Handedness was confirmed using the Edinburgh Handedness Inventory (Oldfield, 1971). Written informed consent was obtained prior to participation in the study. The experimental protocol was approved by the University of Minnesota Institutional Review Board (STUDY00012759).

### Device and training procedure

Detailed information about the training device, training procedure and motor assessment was published previously (Winter et al., 2024). Briefly, participants practiced a gamified motor task using a custom-made wrist-robotic device (Error! Reference source not found.**A)**. The motor task, a classic single player virtual car racing game, consisted of a 2 degree-of-freedom (DoF) task in which wrist flexion/extension and forearm pronation/supination were coupled to the movement of the car on the screen (Error! Reference source not found.**B**). All participants completed 20 physical practice trials of 60 seconds each on day 1 and returned for a *retention* test on day 2, approximately 24 hours after the end of training, to perform five trials of the same gamified exercise.

Participants were randomly allocated to one of three groups (each *n* = 14): 1) dyad practice (*DP*), 2) physical practice only with rest (*PP-rest*), or 3) physical practice only without rest (*PP-no rest*). Each *DP* participant was paired with one other *DP* participant, resulting in seven training dyads. All participants were informed that the learning objective of the gamified exercise was to finish the game as fast as possible, and that they were required to keep the virtual car on the road as much as they could. Dyads were further instructed to cooperatively learn the task with their learning partner and to “get better together by helping each other out”. They alternated between physical and observational practice. Between trials, *DP* participants were encouraged to engage in an inter-trial dialog lasting 30 seconds, in which they could discuss relevant strategies or observations with their practice partner. Participants in the other two groups only practiced physically. Instead of engaging in observational practice, *PP-rest* rested for 90 seconds between trials to mimic the overall practice time of the *DP* group. *PP-no rest* performed all 20 trials consecutively without rest. *PP-no rest*, therefore, had a shorter overall practice period than *DP* and *PP-rest*.

**Figure 1:**
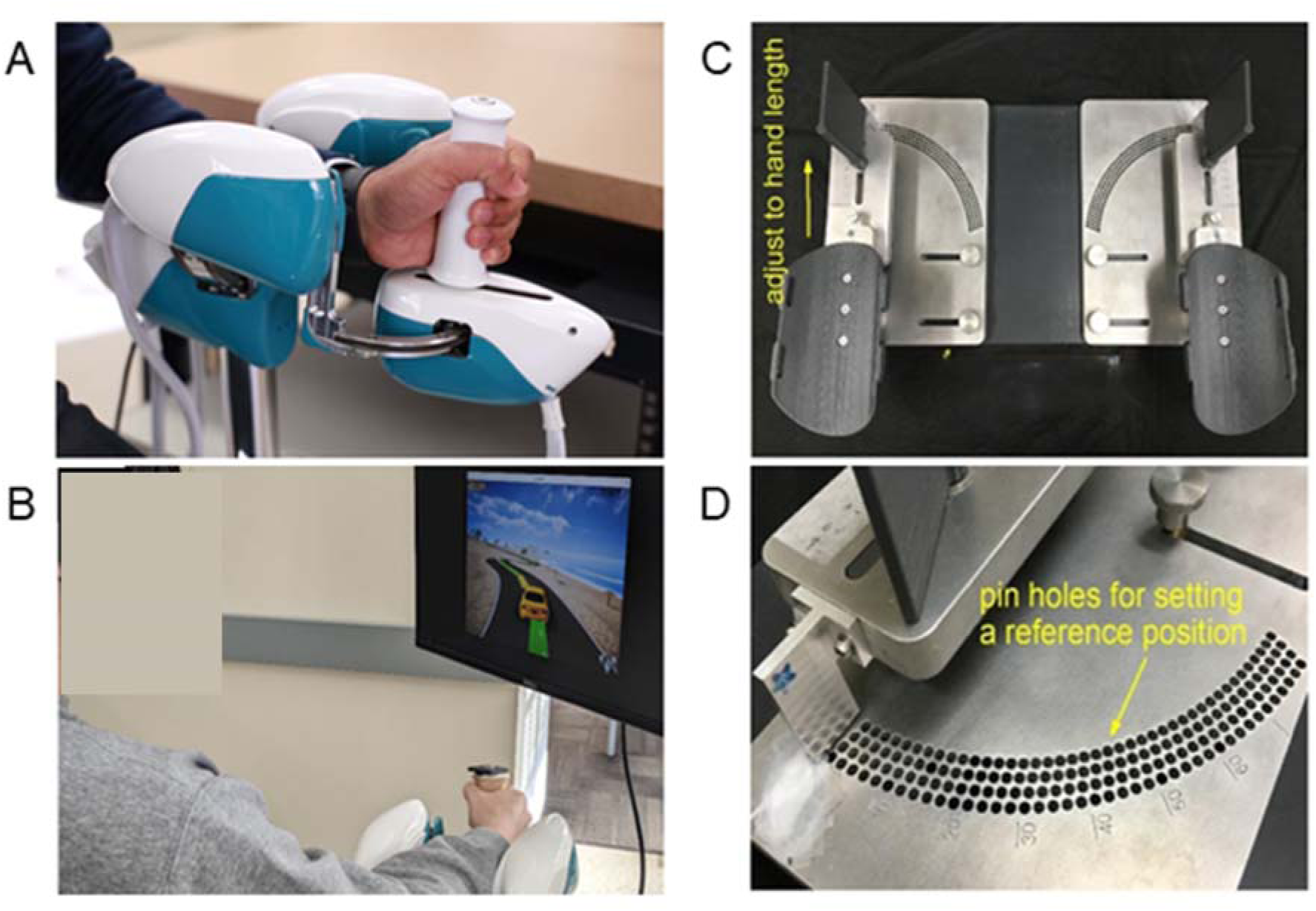
Experimental devices for motor training **(A and B)** and proprioceptive assessment **(C and D)**: Wrist robotic device and gamified exercise (modified from Winter et al., 2024) and wrist manipulandum (modified from Tseng et al., 2018). **A)** Close-up of the wrist robotic device. **B)** Participant during practice of the gamified exercise. The goal was to finish the course as fast as possible. **C)** The wrist manipulandum was used for proprioceptive assessment. Adjustable handles allowed accommodating for different hand size. **D)** A pegboard panel allowed inserting a metal pin into a hole that served as a mechanical stop for a distinct reference position.

### Evaluation of proprioceptive performance improvement

Wrist joint position sense acuity was measured using a custom-built passive motion manipulandum (Tseng et al., 2018) that rotates the wrist in the medio-lateral plane (flexion and extension) (Error! Reference source not found.**C**). An optical encoder with a spatial resolution of 0.036° (U.S. Digital H6), housed at the rotating axis of the lever arm, recorded angular position of the wrist at a sampling rate of 42 Hz. To allow for the testing of distinct joint positions across the joint range of motion, the device has a pegboard with holes in a semicircular arrangement. By inserting a metal pin, researchers could select a precise standard position from 140 different positions across 0° to 70° flexion in 0.5° increments (Error! Reference source not found.**D**). During testing, the participants’ forearm was supported and a handle placed in the participants’ palm.

Wrist joint position sense acuity was measured before the motor task training on day 1 (*baseline*) and after the five motor retention trials on day 2 (*retention*). During the proprioceptive assessment, participants wore opaque glasses to block visual cues and wore headphones playing pink noise to block auditory cues. The researcher manually moved the participant’s hand from a neutral position to one of two positions either at 10° wrist flexion (*standard*) or a flexed position that was always <10° (*comparison)*.

This position was held for 2 seconds, then the wrist was rotated back to the starting position. Subsequently, the hand was moved to a different position (see Error! Reference source not found.**A** for schematic demonstration). Presentation order of the standard and comparison stimuli was randomized. The handle was rotated at a velocity of 6 degrees/second.

After both movements were complete, the participant indicated verbally which movement was further from the starting position (*first* or *second*). Based on the response under a forced-choice paradigm, an adaptive psi-marginal algorithm used the correctness of the previous response and the previous stimulus size to select the next comparison stimulus (Prins, 2013). Each test was composed of 20 trials. One break was provided after 10 trials. Error! Reference source not found.**B** displays exemplary results from one participant over 20 trials.

After the completion of the test, a logistic-Weibull function was fitted to the stimulus size difference between the comparison and reference position and the corresponding percentage of correct responses (Error! Reference source not found.**C)**. Based on the fitted function, two outcome measures were calculated: (1) *Just-noticeable-difference (JND)* threshold and (2) *Uncertainty*. The *JND* threshold is a measure of position sense bias, i.e., the systematic error when perceiving joint position sense. *JND* threshold is defined as the stimulus size at which the participant could correctly perceive their wrist position with 75% accuracy. Smaller *JND* thresholds indicate higher position sense acuity. The *Uncertainty* is measured as the slope of the fitted function at the *JND* threshold. *Uncertainty* is a measure of position sense precision, i.e., the random error. It expresses the consistency of a participant’s response when repeatedly experiencing a specific joint position. A larger value of *Uncertainty* indicates a higher position sense acuity.

**Figure 2.**
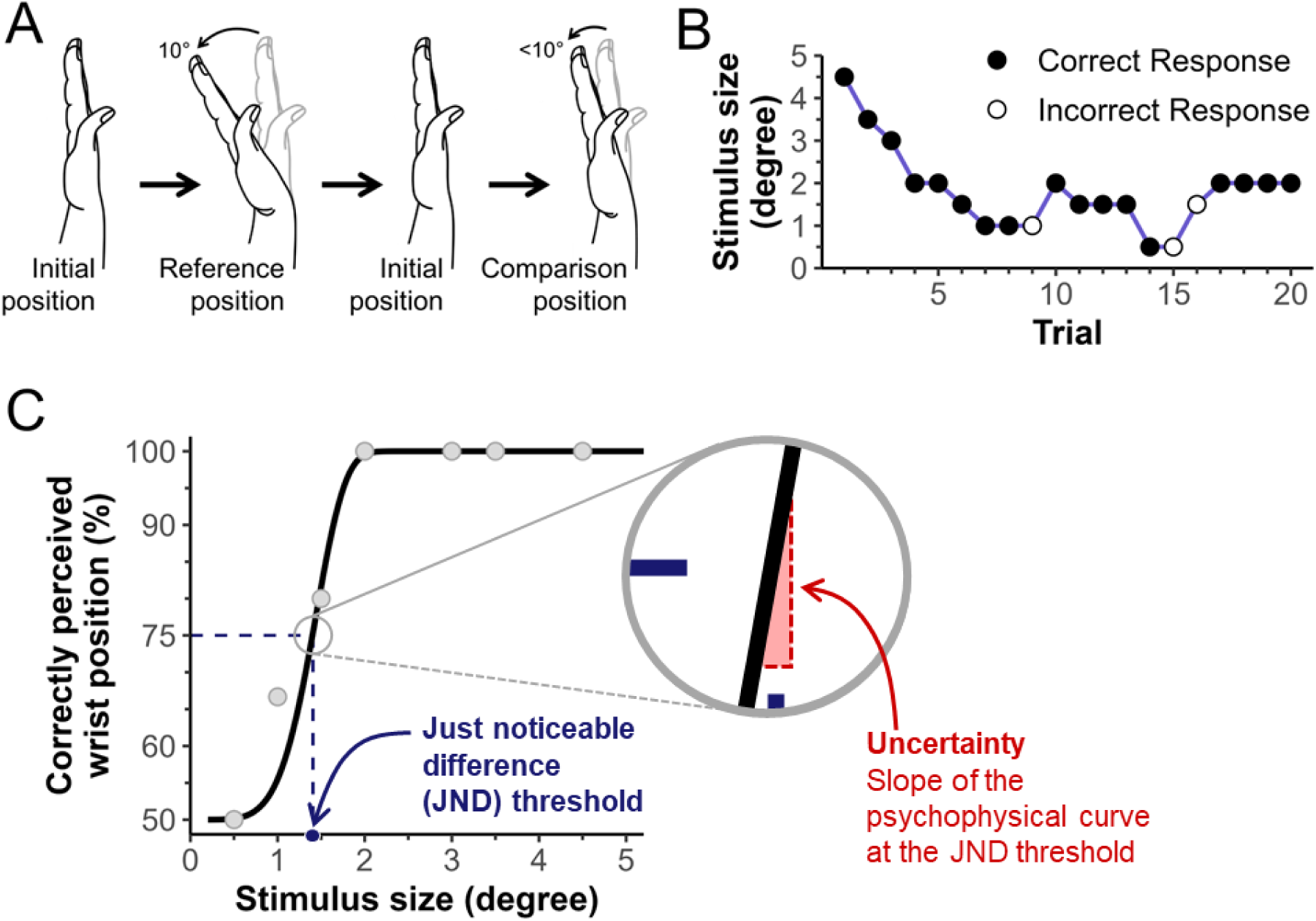
Proprioceptive testing. **A)** Illustration of a single trial of wrist position sense testing. **B)** Exemplary data of participant responses at each stimulus size over 20 trials. **C)** Percentage of correct responses at the presented stimulus sizes. Dots are the stimulus sizes presented in *B*. A logistic-Weibull function is fitted to the data. *JND* threshold is the stimulus size where the participant was predicted to have a 75% correct response rate based on the fitted function. The steepness of the curve at the *JND* threshold is the *Uncertainty*

The reason for assessing proprioception at baseline and retention, rather than at baseline and immediately post-training was two-fold: First, we wanted to avoid mental fatigue for participants on the first day of the study. Second, previous research shows that somatosensory training effects remain stable for several days after training (Cuppone et al., 2018). Therefore, participants were assessed after a 24 hour retention period, which ensured that the best possible data was available.

### Statistical analysis

The distribution for all variables were examined for normality using Shapiro-Wilk’s test of normality. *JND* threshold was normally distributed for all groups but *Uncertainty* was not normally distributed in the *DP* group. Consequently, parametric analyses were performed for *JND* threshold, and non-parametric analyses were performed for *Uncertainty.* A simple mixed method analysis of variance (ANOVA) was performed to identify main effects of time (*baseline, retention*) and group (*DP, PP-rest, PP-no rest*). Post-hoc *t*-tests were used to identify individual group differences from *baseline* to *retention*. Kruskal-Wallis H tests were performed to determine between-group differences in *Uncertainty* at *baseline* and *retention*. Post-hoc Mann-Whitney U tests were used to identify individual group differences from *baseline* to *retention*. Hedge’s g effect sizes were calculated for within-group and between-group differences for both *JND* threshold and *Uncertainty*. The significance level for all tests was set to α = 0.05. All statistical analyses were conducted using MATLAB version 2023b (MathWorks, Inc., Natick, MA).

## Results

The associated motor performance data are published in detail elsewhere (Winter et al., 2024). Briefly, all groups improved their motor performance from *baseline* to the end of training, while dyads showed larger performance gains than both *PP* groups. Motor gains remained larger for dyads after a 24-h *retention* period, compared to the two *PP* groups.

### Practice related changes in position sense bias

Error! Reference source not found. illustrates the *JND* threshold at *baseline* and *retention* for the three groups: *PP-no rest*, *PP-rest*, and *DP. Baseline* and *retention* mean *JND* thresholds were 1.4° and 1.1° for the *PP-no rest* group, 1.6° and 1.3° for the *PP-rest* group, and 1.4° and 1.3° for the *DP* group, respectively (see **Table 1**, Error! Reference source not found.**A**). Simple mixed method ANOVA revealed significant main effect of time (*F_(1,39)_*=10.06; *p*=0.003, η²=0.21) and no group effect (*F_(2,39)_*=0.53; *p*=0.59, η²=0.03). For the *PP-no rest* group, position sense bias at *retention* improved by 20% compared to *baseline* (*t*(13)=2.44; *p*= 0.03, *p* = 0.03, Hedge’s *g* = 0.70). Ten of the 14 *PP-no rest* participants demonstrated higher proprioceptive acuity at *retention* than at *baseline* (Error! Reference source not found.**B**). That is, 71% of participants showed improvements in wrist joint position sense bias 24 hours after practicing the gamified exercise. Only considering responders, participants in the *PP-no rest* group improved their performance by 31%, on average. In contrast, differences in *JND* threshold between *baseline* and *retention* were not statistically significant for the *PP-rest* and the *DP* group. However, it is noteworthy that 10 *PP-rest* participants (71%) and eight *DP* participants (57%) showed improvements in *JND* threshold at *retention*. Considering only individuals who improved their performance, responders showed a reduction in *JND* threshold by 30% and 26% for the *PP-rest* and the *DP* groups, respectively. Overall, 28/42 (66%) of participants across all three practice groups exhibited a reduction in JND threshold at retention relative to baseline.

**Table 1.**
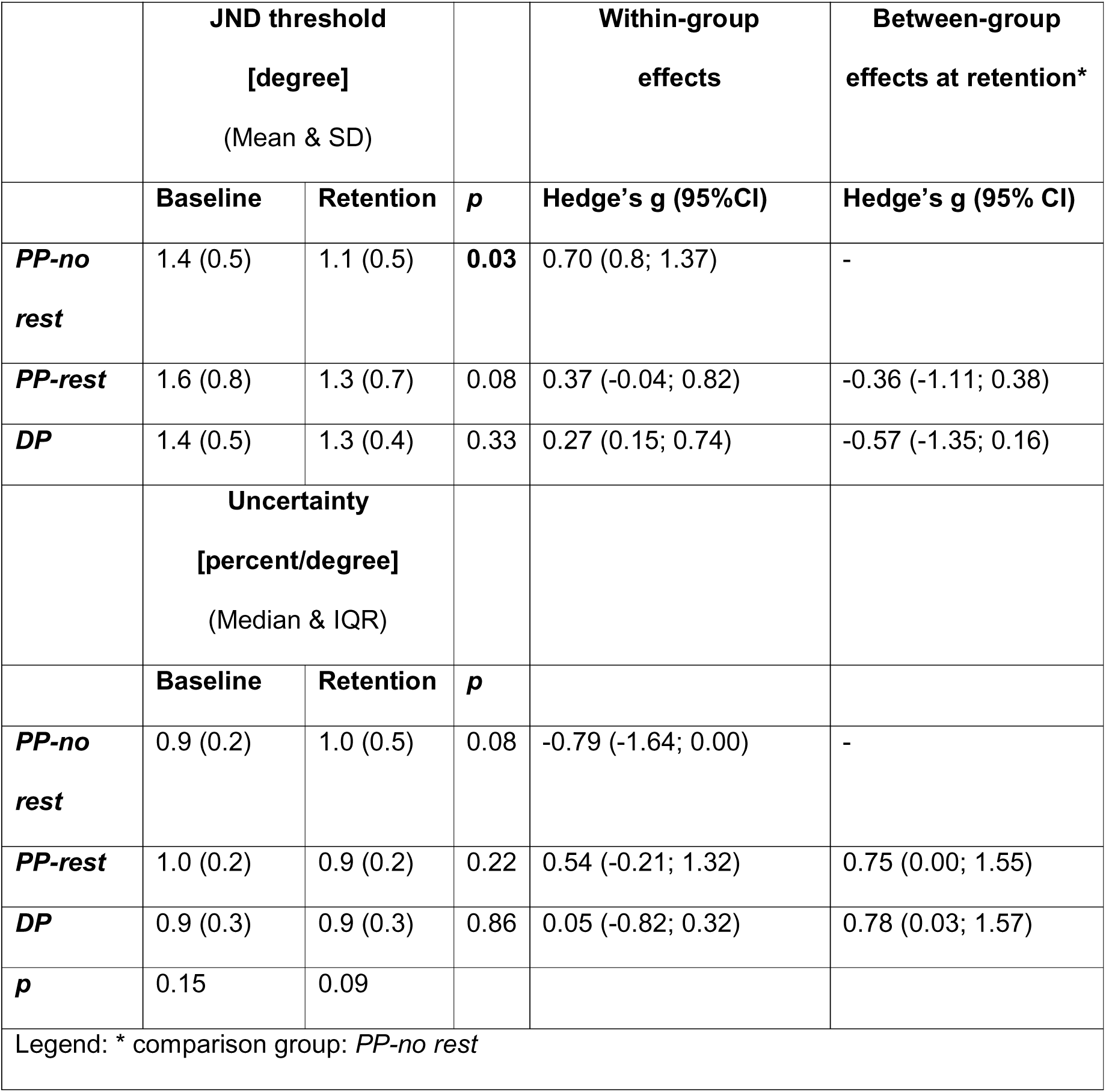
Summary statistics of *JND* threshold and *Uncertainty* at *baseline* and *retention* for each practice group. Effect sizes for within-group and between-group effects are expressed by Hedge’s g. Values in parentheses represent the standard deviation (SD) for *JND* threshold, and interquartile ranges (IQR) for *Uncertainty*. *PP-no rest*: physical practice group with no rest between trials; *PP-rest*: physical practice group with rest between trials; *DP*: dyad practice group.

|  | **JND threshold**<br>**[degree]**<br>(Mean & SD) |  |  | **Within-group**<br>**effects** | **Between-group**<br>**effects at retention\*** |
| --- | --- | --- | --- | --- | --- |
|  | **Baseline** | **Retention** | ***p*** | **Hedge's *g* (95%CI)** | **Hedge's *g* (95% CI)** |
| ***PP-no rest*** | 1.4 (0.5) | 1.1 (0.5) | **0.03** | 0.70 (0.8; 1.37) | - |
| ***PP-rest*** | 1.6 (0.8) | 1.3 (0.7) | 0.08 | 0.37 (-0.04; 0.82) | -0.36 (-1.11; 0.38) |
| ***DP*** | 1.4 (0.5) | 1.3 (0.4) | 0.33 | 0.27 (0.15; 0.74) | -0.57 (-1.35; 0.16) |
|  | **Uncertainty**<br>**[percent/degree]**<br>(Median & IQR) |  |  |  |  |
|  | **Baseline** | **Retention** | ***p*** |  |  |
| ***PP-no rest*** | 0.9 (0.2) | 1.0 (0.5) | 0.08 | -0.79 (-1.64; 0.00) | - |
| ***PP-rest*** | 1.0 (0.2) | 0.9 (0.2) | 0.22 | 0.54 (-0.21; 1.32) | 0.75 (0.00; 1.55) |
| ***DP*** | 0.9 (0.3) | 0.9 (0.3) | 0.86 | 0.05 (-0.82; 0.32) | 0.78 (0.03; 1.57) |
| ***p*** | 0.15 | 0.09 |  |  |  |
| Legend: * comparison group: *PP-no rest* |  |  |  |  |  |

### Practice related changes in position sense precision

Error! Reference source not found. illustrates the *Uncertainty* measured by the steepness of the log-Weibull function at *baseline* and *retention* for each group. At *baseline* and *retention*, the median *Uncertainty* was 0.9 and 1 percent/degree for the *PP-no rest* group, 1.0 and 0.9 percent/degree in the *PP-rest* group, and 0.9 percent/degree for both *baseline* and *retention* in the *DP* group, respectively (**Table 1**, Error! Reference source not found.**A**). Nine *PP-no rest* participants (64%), five *PP-rest* participants (36%), and seven *DP* participants (50%) demonstrated smaller *Uncertainty* at *retention* than at *baseline* (Error! Reference source not found.**B**). However, changes were not statistically significant for any of the within-and between-group comparisons.

**Figure 3.**
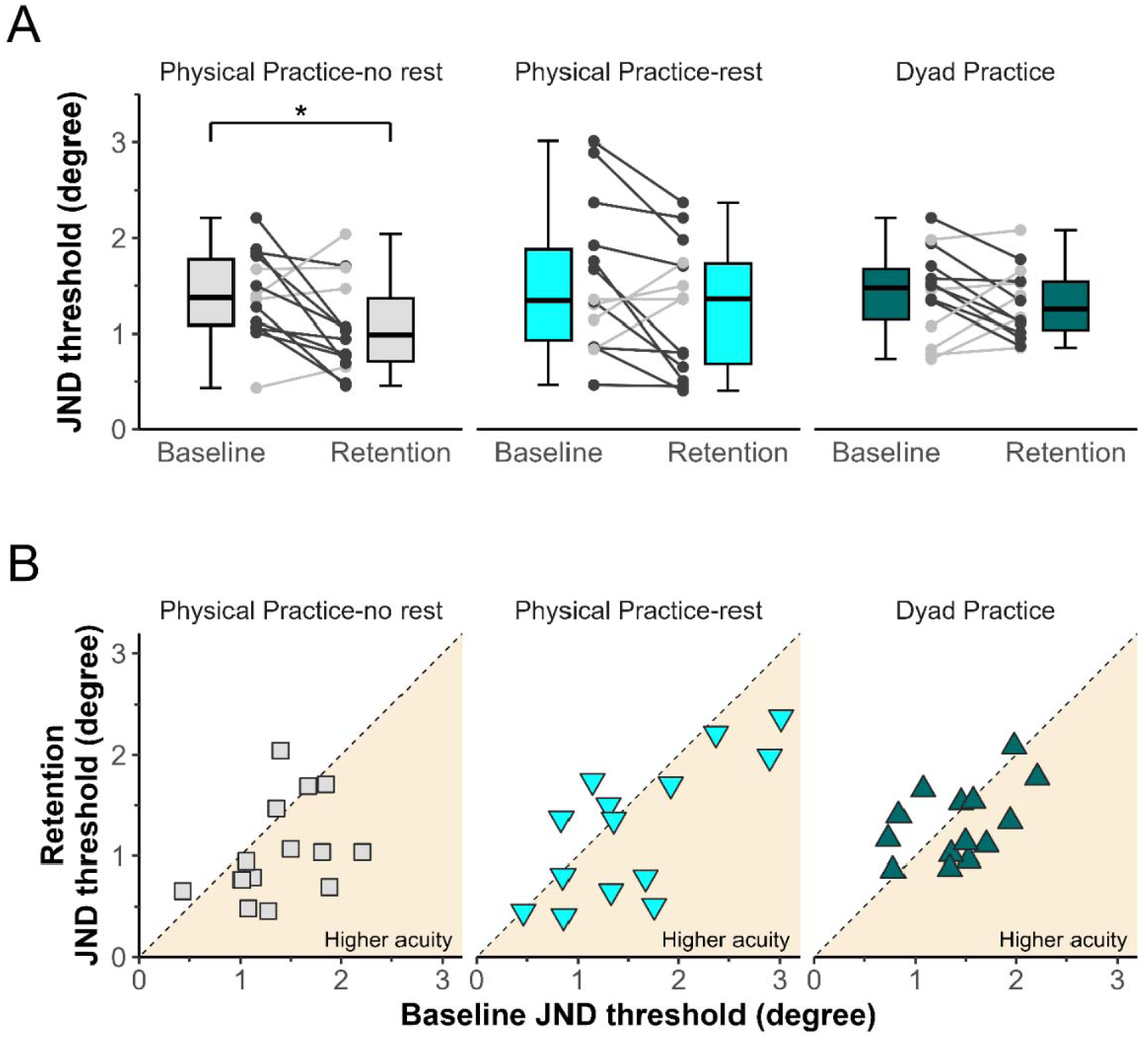
*JND* wrist position sense thresholds at *baseline* and *retention*. Individual participant performance is shown as points on both A and B. **A)** *JND* threshold at *baseline* and 24-hr *retention* for each group. Significant between-group differences at *retention* are indicated by an asterisk. Whiskers represent the 5th and 95th percentiles. Participants are represented as circles and connected by lines to indicate individual change from *baseline* to *retention*. Dark grey points indicate higher proprioceptive acuity at *retention* than *baseline,* whereas light grey points represent lower acuity after the intervention. **B)** Comparison of *JND* threshold at *baseline* and at *retention* for each participant in each group. The dashed diagonal line represents the *line of equality*. Participants with improved *JND* threshold at *retention* (higher acuity) fall below the line of equality.

**Figure 4.**
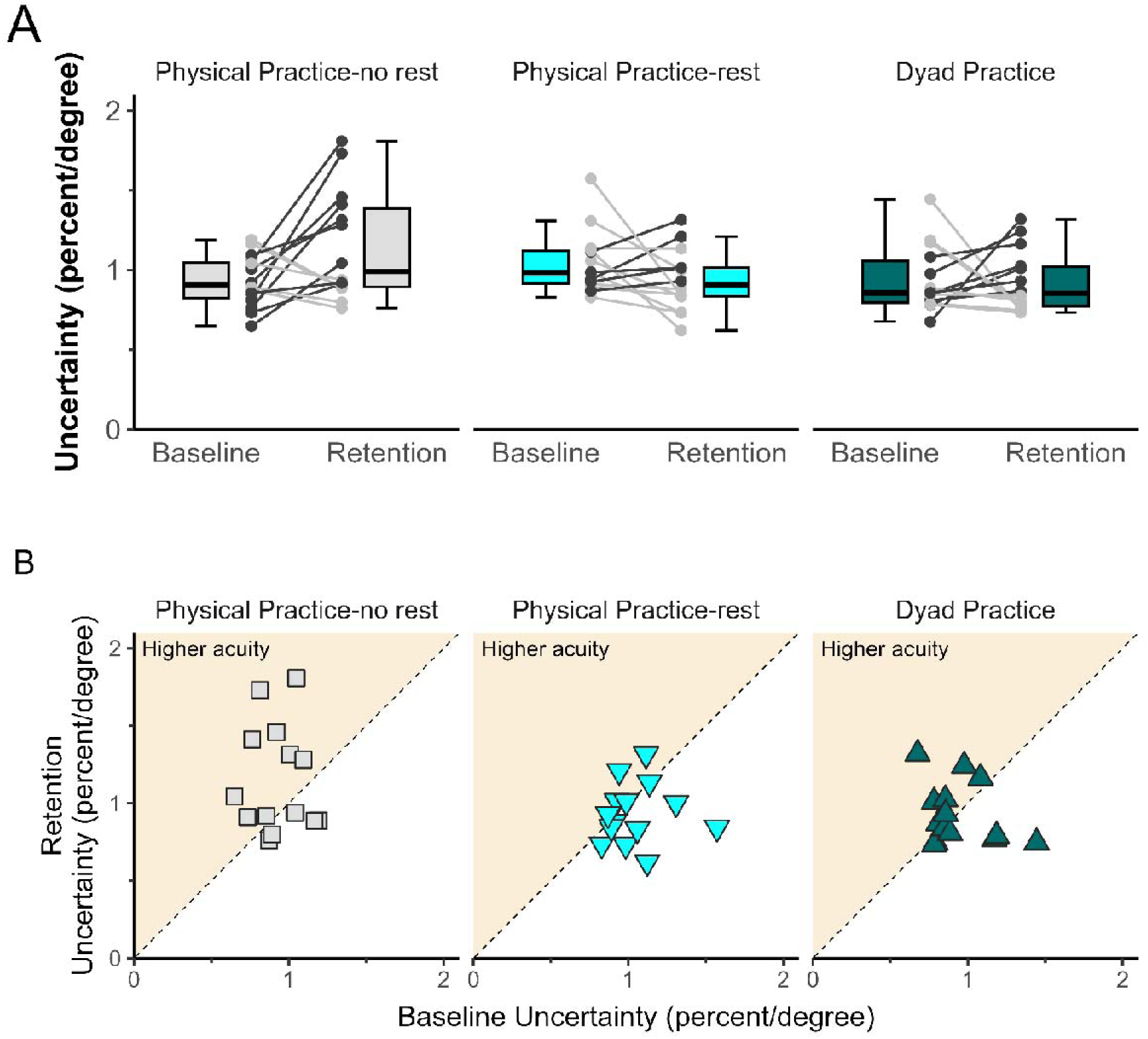
Wrist position sense *Uncertainty* at *baseline* and *retention*. **A)** *Uncertainty* at *baseline* and 24-hr *retention* for each group. Whiskers represent the 5th and 95th percentiles. Participants are represented as circles and connected by lines, which show their change from *baseline* to *retention*. Dark grey points indicate higher proprioceptive acuity at *retention* than *baseline*. Light grey points represent lower wrist proprioceptive acuity at *retention* than *baseline*. **B)** Comparison of *Uncertainty* at *baseline* and at *retention* for each participant in each group. The dashed line represents the *line of equality* (diagonal). Participants with improved *Uncertainty* at *retention* (higher acuity) are above the line of equality.

## Discussion

To our knowledge, this is the first study to assess the effects of dyad practice on proprioceptive performance. The main results were threefold: First, on a group level, only the physical practice group with no rest interval exhibited a significantly lower mean JND position sense threshold 24 hours after training. Second, across the three practice groups, the majority of participants (66%) showed reduced position sense *JND* thresholds at retention. Third, *Uncertainty,* representing a measure of precision or random error when judging position differences across repeated trials, did not differ across practice groups.

### Dyad sensorimotor practice did not lead to superior proprioceptive gains

The study findings show that with identical amounts of physical practice in all groups, additional observational practice does not lead to superior proprioceptive acuity in dyads. To understand why dyad practice during the acquisition of complex motor skills yields additional benefits in motor performance and learning economy, but has no discernible influence on proprioceptive learning, it is useful to consider the learning mechanisms and the underlying neural networks associated with motor and proprioceptive learning. Empirical research in human and non-human primates suggests that complex motor skill learning typically involves two distinct learning mechanisms: implicit (unconscious) and explicit (conscious) learning. Various forms of motor learning such as adaptation, conditioning and skill acquisition represent implicit or procedural learning and activates distinct brain areas including the cerebellum, basal ganglia, sensorimotor cortex, amygdala (Squire & Zola, 1996; Yang & Li, 2012). In contrast, explicit learning and specifically observational learning activates networks in the medial temporal lobe, hippocampus and lateral prefrontal cortex (Kang et al., 2020; Squire & Zola, 1996). Dyad practice engages both explicit and implicit learning networks. That is, the physical training portion during dyad practice relies on the same implicit learning networks that are known to be active during motor learning.

Consequently, the dyad learners in this study, who trained the same amount as the other two physical practice groups, showed similar effects on proprioceptive function. The additional activation of explicit learning networks in the dyad learning group likely did not enhance short-term somatosensory memory formation that would become manifest as higher position sense acuity.

### Higher physical practice intensity benefits proprioceptive performance

The more consistent and larger proprioceptive gains in the *PP-no rest* group indicate that participants in this group benefitted from more intense, non-interupted physical practice, compared to the *PP-rest* and the *DP* groups. That is, despite having practiced the same amount, the *PP-no rest* group had a higher training intensity since participants in this group did not rest in between trials. It has long been established that spaced practice (i.e., physical practice that is distributed over a longer period of time) leads to better motor outcomes than when individuals perform all training trials continuously (Donovan & Radosevich, 1999; Shea et al., 2000). Our results suggest, however, that higher intensity had a positive effect on proprioceptive performance.

While consensus on optimal dosage of proprioceptive training is lacking, higher training intensity may be required in dyad practice to elicit larger proprioceptive gains. Indeed, it would be misleading to say that *DP* did not lead to proprioceptive gains, when in fact, a majority of *DP* participants did improve their position sense bias, measured by *JND* threshold and half of *DP* participants improved their position sense precision, measured by *Uncertainty*. However, it can be concluded that the additional observational practice in the *DP* group was unable to elicit additional proprioceptive improvement, compared to the *PP-rest* group, which practiced at the same level of physical intensity as the *DP* group. In summary, our results show that despite the close relationship between proprioceptive and motor learning, the unique explicit learning opportunities afforded by dyad practice do not benefit proprioceptive performance.

### Limitations

Participants trained a 2-DoF motor task (wrist flexion/extension; forearm supination/pronation), but proprioceptive performance was only assessed for the wrist DoF. Due to the nature of the task, participants were not constrained when and what DoF to use during practice. Therefore, it is possible that different amounts of wrist vs. forearm use across participants could have induced inter-individual differences in our measures of proprioceptive acuity. However, we have no indication that the dyad group was biased, that is, its participants systematically used a different DoF.

Participants’ wrist proprioception was not assessed directly after training but rather, after a 24-hour retention period. It is noteworthy that proprioceptive assessments require active focus by the participant. We aimed to avoid mental fatigue, due to the length of the experiment, that may have negatively affected the data quality of the second proprioceptive assessment. Simultaneously, there is compelling evidence that somatosensory training effects remain stable over several days after the end of training (Cuppone et al., 2018). That is, it is unlikely that training gains were lost during the retention period.

## Conclusion

Dyad practice is effective in improving motor performance by affording unique learning opportunities that benefit learners during complex motor task acquisition. The additional interspersed observational practice allows individuals to develop a cognitive framework of the task, therefore leading to larger practice effects than physical practice only. However, dyad practice is not superior in improving proprioceptive performance, compared to physical practice alone. Intensive physical practice without rest intervals between trials resulted in the largest gains in proprioceptive performance. Therefore, more intense practice may be required for proprioceptive performance changes, than was present in the *DP* and *PP-rest* groups.

The lack of superior proprioceptive performance in the *DP* group suggests that explicit knowledge of the appropriate movement pattern required to perform the motor task does not directly translate into larger gains in proprioceptive acuity. While the explicit information gathered through observational practice clearly benefits motor performance (Winter et al., 2024), the present results indicate that additional observational practice likely does not excite relevant networks involved in the formation of proprioceptive memory.

## Funding

This research was supported by U.S. National Science Foundation IIP 1919036 PFI-TT: A robotic device for the physical therapy of wrist/hand function award to JK. Role of the funding source: The funders played no role in the design, conduct, or reporting of this study.

## Data Availability

All data produced in the present study are available upon reasonable request to the authors.

## Abbreviations

DoF: Degree-of-freedom
DP: Dyad practice
JND: Just-Noticeable-Difference
PP-no rest: Physical practice only without rest
PP-rest: Physical practice only with rest

## Acknowledgements

We thank Naveen Elangovan, Daniel Sousa and Reuben Gardos Reid for their technical support and expertise with the wrist robot, and Shima Amini and Jason Kang for their help with data collection. We are thankful to the volunteers for their participation in the study.

## Notes

### Competing Interest Statement

The authors have declared no competing interest.

### Funding Statement

This study did not receive any funding.

### Author Declarations

Institutional Review Board of the University of Minnesota gave ethical approval for this work.

## References

Aman, J. E., Elangovan, N., Yeh, I.-L., & Konczak, J. (2015). The effectiveness of proprioceptive training for improving motor function: A systematic review. Frontiers in Human Neuroscience, 8(JAN). 10.3389/fnhum.2014.01075

Arthur, W., Day, E. A., Bennett, W., McNelly, T. L., & Jordan, J. A. (1997). Dyadic versus individual training protocols: Loss and reacquisition of a complex skill. Journal of Applied Psychology, 82(5), 783–791. 10.1037/0021-9010.82.5.783

Bjerrum, A. S., Eika, B., Charles, P., & Hilberg, O. (2014). Dyad practice is efficient practice: A randomised bronchoscopy simulation study. Medical Education, 48(7), 705–712. 10.1111/medu.12398

Borich, M. R., Brodie, S. M., Gray, W. A., Ionta, S., & Boyd, L. A. (2015). Understanding the role of the primary somatosensory cortex: Opportunities for rehabilitation. Neuropsychologia, 79, 246–255. 10.1016/j.neuropsychologia.2015.07.007

Cuppone, A. V., Semprini, M., & Konczak, J. (2018). Consolidation of human somatosensory memory during motor learning. Behavioural Brain Research, 347, 184–192. 10.1016/j.bbr.2018.03.013

Darainy, M., Manning, T. F., & Ostry, D. J. (2023). Disruption of somatosensory cortex impairs motor learning and retention. Journal of Neurophysiology, 130(6), 1521–1528. 10.1152/jn.00231.2023

Donovan, J. J., & Radosevich, D. J. (1999). A meta-analytic review of the distribution of practice effect: Now you see it, now you don’t. Journal of Applied Psychology, 84(5), 795–805.

Elangovan, N., Cappello, L., Masia, L., Aman, J., & Konczak, J. (2017). A robot-aided visuo-motor training that improves proprioception and spatial accuracy of untrained movement. Scientific Reports, 7(1), 17054. 10.1038/s41598-017-16704-8

Elangovan, N., Tuite, P. J., & Konczak, J. (2018). Somatosensory Training Improves Proprioception and Untrained Motor Function in Parkinson’s Disease. Frontiers in Neurology, 9(December). 10.3389/fneur.2018.01053

Elangovan, N., Yeh, I. L., Holst-Wolf, J., & Konczak, J. (2019). A robot-assisted sensorimotor training program can improve proprioception and motor function in stroke survivors. IEEE International Conference on Rehabilitation Robotics, 2019*-June*, 660–664. 10.1109/ICORR.2019.8779409

Granados, C., & Wulf, G. (2007). Enhancing motor learning through dyad practice: Contributions of observation and dialogue. Research Quarterly for Exercise and Sport, 78(3), 197–203. 10.1080/02701367.2007.10599417

Kang, W., Pineda Hernández, S., & Mei, J. (2020). Neural Mechanisms of Observational Learning: A Neural Working Model. Frontiers in Human Neuroscience, 14, 609312. 10.3389/fnhum.2020.609312

Oldfield, R. C. (1971). The assessment and analysis of handedness: The Edinburgh inventory. Neuropsychologia, 9(1), 97–113.

Ostry, D. J., Darainy, M., Mattar, A. A. G., Wong, J., & Gribble, P. L. (2010). Somatosensory plasticity and motor learning. Journal of Neuroscience, 30(15), 5384–5393. 10.1523/JNEUROSCI.4571-09.2010

Ostry, D. J., & Gribble, P. L. (2016). Sensory Plasticity in Human Motor Learning. Trends in Neurosciences, 39(2), 114–123. 10.1016/j.tins.2015.12.006

Panzer, S., Haab, T., Massing, M., Pfeifer, C., & Shea, C. H. (2019). Dyad training protocols and the development of a motor sequence representation. Acta Psychologica, 201(August), 102947. 10.1016/j.actpsy.2019.102947

Prins, N. (2013). The psi-marginal adaptive method: How to give nuisance parameters the attention they deserve (no more, no less). Journal of Vision, 13(7), Article 7. 10.1167/13.7.3

Seo, H. G., Yun, S. J., Farrens, A. J., Johnson, C. A., & Reinkensmeyer, D. J. (2023). A Systematic Review of the Learning Dynamics of Proprioception Training: Specificity, Acquisition, Retention, and Transfer. Neurorehabilitation and Neural Repair, 37(10), 744–757. 10.1177/15459683231207354

Shea, C. H., Lai, Q., Black, C., & Park, J.-H. (2000). Spacing practice sessions across days benefits the learning of motor skills. Human Movement Science, 19(5), 737–760. 10.1016/S0167-9457(00)00021-X

Shea, C. H., Whitacre, C., & Wulf, G. (1999). Enhancing training efficiency and effectiveness through the use of dyad training. Journal of Motor Behavior, 31(2), 119–125. 10.1080/00222899909600983

Shebilske, W. L., Jordan, J. A., Goettl, B. P., & Paulus, L. E. (1998). Observation versus hands-on practice of complex skills in dyadic, triadic, and tetradic training-teams. Human Factors, 40(4), 525–540. 10.1518/001872098779649319

Squire, L. R., & Zola, S. M. (1996). Structure and function of declarative and nondeclarative memory systems. Proceedings of the National Academy of Sciences of the United States of America, 93(24), 13515–13522. 10.1073/pnas.93.24.13515

Tolsgaard, M. G., Madsen, M. E., Ringsted, C., Oxlund, B. S., Oldenburg, A., Sorensen, J. L., Ottesen, B., & Tabor, A. (2015). The effect of dyad versus individual simulation-based ultrasound training on skills transfer. Medical Education, 49(3), 286–295. 10.1111/medu.12624

Tseng, Y.-T., Tsai, C.-L., Chen, F.-C., & Konczak, J. (2018). Wrist position sense acuity and its relation to motor dysfunction in children with developmental coordination disorder. Neuroscience Letters, 674, 106–111. 10.1016/j.neulet.2018.03.031

Vahdat, S., Darainy, M., Milner, T. E., & Ostry, D. J. (2011). Functionally specific changes in resting-state sensorimotor networks after motor learning. The Journal of Neuroscience: The Official Journal of the Society for Neuroscience, 31(47), 16907–16915. 10.1523/JNEUROSCI.2737-11.2011

Winter, L. V., Huang, Q., Sertic, J. V. L., & Konczak, J. (2022). The Effectiveness of Proprioceptive Training for Improving Motor Performance and Motor Dysfunction: A Systematic Review. Frontiers in Rehabilitation Sciences, 3(April). 10.3389/fresc.2022.830166

Winter, L. V., Panzer, S., & Konczak, J. (2024). Dyad motor learning in a wrist-robotic environment: Learning together is better than learning alone. Human Movement Science, 93, 103172. 10.1016/j.humov.2023.103172

Yang, J., & Li, P. (2012). Brain networks of explicit and implicit learning. PloS One, 7(8), e42993. 10.1371/journal.pone.0042993

